# Germline mosaicism of a missense variant in *KCNC2* in a multiplex family with autism and epilepsy

**DOI:** 10.1101/2021.12.06.21264306

**Authors:** Elvisa Mehinovic, Teddi Gray, Meghan Campbell, Jenny Ekholm, Aaron Wenger, William Rowell, Ari Grudo, Jane Grimwood, Jonas Korlach, Christina Gurnett, John N. Constantino, Tychele N. Turner

## Abstract

Currently, protein-coding *de novo* variants and large copy number variants have been identified as important for ∼30% of individuals with autism. One approach to identify relevant variation in individuals who lack these types of events is by utilizing newer genomic technologies. In this study, highly accurate PacBio HiFi long-read sequencing was applied to a family with autism, treatment-refractory epilepsy, cognitive impairment, and mild dysmorphic features (two affected female full siblings, parents, and one unaffected sibling) with no known clinical variant. From our long-read sequencing data, a *de novo* missense variant in the *KCNC2* gene (encodes Kv3.2 protein) was identified in both affected children. This variant was phased to the paternal chromosome of origin and is likely a germline mosaic. *In silico* assessment of the variant revealed it was in the top 0.05% of all conserved bases in the genome, and was predicted damaging by Polyphen2, MutationTaster, and SIFT. It was not present in any controls from public genome databases nor in a joint-call set we generated across 49 individuals with publicly available PacBio HiFi data. This specific missense mutation (Val473Ala) has been shown in both an ortholog and paralog of Kv3.2 to accelerate current decay, shift the voltage dependence of activation, and prevent the channel from entering a long-lasting open state. Seven additional missense mutations have been identified in other individuals with neurodevelopmental disorders (p = 1.03 × 10^−5^). *KCNC2* is most highly expressed in the brain; in particular, in the thalamus and is enriched in GABAergic neurons. Long-read sequencing was useful in discovering the relevant variant in this family with autism that had remained a mystery for several years and will potentially have great benefits in the clinic once it is widely available.

## INTRODUCTION

Autism is a complex neurodevelopmental disorder with a genetic component from both common and rare variation. Common variation contributes to ∼50% of autism [Gaugler and others 2014] and individuals with autism have excess polygenic risk [Weiner and others 2017]. In terms of rare variation, ∼30% of all cases can be explained by *de novo* copy number variants and *de novo* single-nucleotide variants or small insertions/deletions that are loss-of-function or severe missense changes [Iossifov and others 2014]. Several other types of rare genetic variants have been implicated in autism including rare inherited variants [Iossifov and others 2015; Krumm and others 2015; Wilfert and others 2021], recessive variants [Doan and others 2019], and *de novo* noncoding putative regulatory variants [An and others 2018; Padhi and others 2021; Turner and others 2017; Turner and others 2016; Zhou and others 2019]. Another important feature of autism is its sex ratio of 4:1 with 80% of all cases being male. There has been a documented excess of rare *de novo* variants and recessive variants in females with autism [Doan and others 2019; Iossifov and others 2014; Jacquemont and others 2014; Levy and others 2011; Neale and others 2012; Sanders and others 2015; Turner and others 2019] and families with multiple affected females with autism have been prioritized for gene discovery [Turner and others 2015].

Recently, long-read sequencing technologies have emerged and enabled access to variation previously intractable with short-read sequencing [Wenger and others 2019]. These technologies are promising for the objective of precision genomics (i.e., complete resolution of all genomic variation in an individual) for precision medicine. Furthermore, family-based studies have been highly successful in identifying *de novo* variants and long-read sequencing coupled with family-based analyses should prove especially fruitful for gene discovery in families with no known genetic cause.

This study was focused on a family with two females with autism and epilepsy, their unaffected sibling, and their unaffected parents. This family had previously undergone whole-exome sequencing and array analyses in the clinic with no identification of relevant genomic variation. Since there were two affected females in the family and they also shared additional phenotypes including epilepsy, we hypothesized that long-read sequencing technologies would uncover the relevant genomic variation in this family. Pacific Biosciences long-read sequencing, 10x Genomics sequencing, and Bionano optical mapping were applied for each family member and from this analysis a missense *de novo* variant in the *KCNC2* gene (encodes the Kv3.2 potassium channel) was identified in both females with autism but not in their unaffected sibling. This variant arose on the paternal chromosome in both individuals and is likely to be a germline mosaic event. There are multiple lines of evidence that support the importance of this variant for the family in this study and provides an example of the use of precision genomics, via long-read sequencing, in autism.

## MATERIALS AND METHODS

### Family enrollment in the study

The Washington University in St. Louis Institutional Review Board approved this research under IRB ID #202002147. The Washington University Federal Wide Assurance (FWA) number is FWA00002284, the Washington University IRB is IRB00009237, and the Washington University Protocol Adherence Review Committee (PARC) IRB is IRB00005594. The family (PB.100) was consented using approved Consent and Assent forms (approved in IRB ID #202002147) during a visit to the Washington University in St. Louis Child Psychiatry Clinic. During this visit a total of 2 × 25 ml tubes of blood were collected from each of the five family members (parents, two female children with autism and epilepsy, and one unaffected child). One 25 ml tube of blood was taken to the McDonnell Genome Institute for high molecular weight DNA extraction through their core services and the other was taken to the Washington University in St. Louis Genome Engineering and iPSC center for PBMC storage through their core services.

### Genomic technologies

Both the 10x Genomics and the Bionano technologies were applied to DNA from all family members at the McDonnell Genome Institute through their core services following standard protocols. The PacBio HiFi sequencing from DNA in all family members was performed at the HudsonAlpha Genome Sequencing Center through a PacBio SMRT grant.

### Analysis of 10x Genomics genome data

Longranger 2.2.2 (https://support.10xgenomics.com/genome-exome/software/downloads/latest) was run on everyone using the longranger wgs command, fastq as input, FreeBayes [Garrison E 2012] as the variant caller, and the 10x GRCh38-2.1.0 reference genome data. The output loupe files were visualized in the Loupe 2.1.2 browser (https://support.10xgenomics.com/genome-exome/software/downloads/latest). Summary statistics and variant calls were assessed in the browser.

### Analysis of PacBio HiFi genome data in family PB.100

Each CCS fastq file was aligned to build 38 of the human genome (GRCh38_full_analysis_set_plus_decoy_hla.fa) using pbmm2 (https://github.com/PacificBiosciences/pbmm2) version 1.3.0 align. Structural variants were called using pbsv (https://github.com/PacificBiosciences/pbsv) version 2.3.0. SNVs/indel GVCFs were called using DeepVariant [Poplin and others 2018] version 1.0.0 and the GVCFs were joint genotyped using GLNexus version 1.2.7 [Yun and others 2020]. Post-calling, PLINK [Purcell and others 2007] was run to confirm all family relationships as correct. Exomiser [Robinson and others 2014] version 12.1.0 was run on the joint-genotyped vcf file using the HPO term HP:0000729. Two different assemblers (HiCanu [Canu version 2.0] [Nurk and others 2020] and Hifiasm [Cheng and others 2021] version 0.13-r307) were utilized to generate *de novo* assemblies for each individual.

### Analysis of PacBio HiFi genome data from the Pangenome project

There were 49 individuals from the HPRC with publicly available PacBio HiFi data. Individual CCS bam files were downloaded from https://s3-us-west-2.amazonaws.com/human-pangenomics/index.html?prefix=submissions/ and converted to fastq files using PacBio bam2fastq version 1.3.0. Each CCS fastq file was then aligned to build 38 of the human genome (GRCh38_full_analysis_set_plus_decoy_hla.fa) using pbmm2 version 1.3.0 align. Structural variants were called using pbsv version 2.3.0. SNVs/indel GVCFs were called using DeepVariant version 1.0.0 and the GVCFs were joint genotyped using GLNexus version 1.2.7.

### Sanger confirmation of the KCNC2 DNV

Primers (Forward primer: GATCTGTTATGTTCCAGAAGTCGAT, Reverse primer: TAGTGAGCACACACAGTTCAAAAAC) were designed to target the exon (hg38: chr12:75050392-75050761) containing the Val473Ala mutation using Primer3 (https://bioinfo.ut.ee/primer3-0.4.0/). The region was amplified using the Phusion High-Fidelity PCR Master Mix (Thermo-Fisher) and Sanger sequencing was performed at GeneWiz.

### Evolutionary assessment of the KCNC2 exon containing the DNV

We assessed the conservation of the *KCNC2* exon containing the DNV by examining the phyloP [Pollard and others 2010] hg38 100-way scores from http://hgdownload.cse.ucsc.edu/goldenpath/hg38/phyloP100way/hg38.phyloP100way.bw. The scores were converted from a bigwig to a bedgraph (https://hgdownload.cse.ucsc.edu/admin/exe/linux.x86_64/bigWigToBedGraph) and the total number of positions in the file was calculated as well as the number of positions with a score less than the score of the DNV position (phyloP score = 9.29). Of the total 2661892195 positions in the file, 2660671278 had a score lower than the DNV site indicating this site is in the top 0.046% most conserved sites in the genome.

### KCNC2 DNVs identified in other individuals with neurodevelopmental disorders

The literature [Kaplanis and others 2020; Rademacher and others 2020; Satterstrom and others 2020; Vetri and others 2020] was searched for DNVs in individuals with neurodevelopmental disorders and identified 7 additional individuals with DNVs in *KCNC2*. To test whether these DNVs combined with the DNVs in the present study were enriched in the *KCNC2* gene, the denovolyzeR [Samocha and others 2014; Ware and others 2015] and chimpanzee-human [Coe and others 2019] tests were run on the data. The DNVs were plotted on the protein (UniProtKB Q96PR1 [KCNC2_HUMAN]) using PROTTER [Omasits and others 2013].

### Expression assessment of KCNC2

The expression of the KCNC2 gene was examined in different adult tissues in the GTEX database (https://gtexportal.org/home/gene/KCNC2). The expression of this gene across different timepoints and brain regions was visualized in the Brainspan database (https://www.brainspan.org/). Cell-type specific expression was visualized in the Allen Brain Map data (https://portal.brain-map.org) in the whole cortex and hippocampus from an 8-week-old mouse.

### Conservation to Drosophila melanogaster ortholog and Homo sapiens paralog

Initially, we observed a similar amino acid sequence (PVPVIV) in the human paralog Kv1.1 (encoded by *KCNA1* gene) while reading [Peters and others 2011]. In Figure 1 of that paper, the authors compared the *Drosophila melanogaster* ortholog called Shaker to Kv1.1 and found this region was highly conserved and the *specific amino acid change we observe in Kv3.2* was modeled in the Peters et al. paper. It is Kv3.2 Val473Ala, Shaker Val478Ala, and Kv1.1 Val408Ala. To quantitate the conservation formally we performed BLAST [Altschul and others 1990] two sequences (https://blast.ncbi.nlm.nih.gov/Blast.cgi?BLAST_SPEC=blast2seq&LINK_LOC=align2seq&PAGE_TYPE=BlastSearch) comparing Shaker and Kv3.2 as well as comparing Kv3.2 and Kv1.1. Each of these formal tests showed high conservation of the S6 transmembrane domain in which the amino acid change resides.

**Figure 1:**
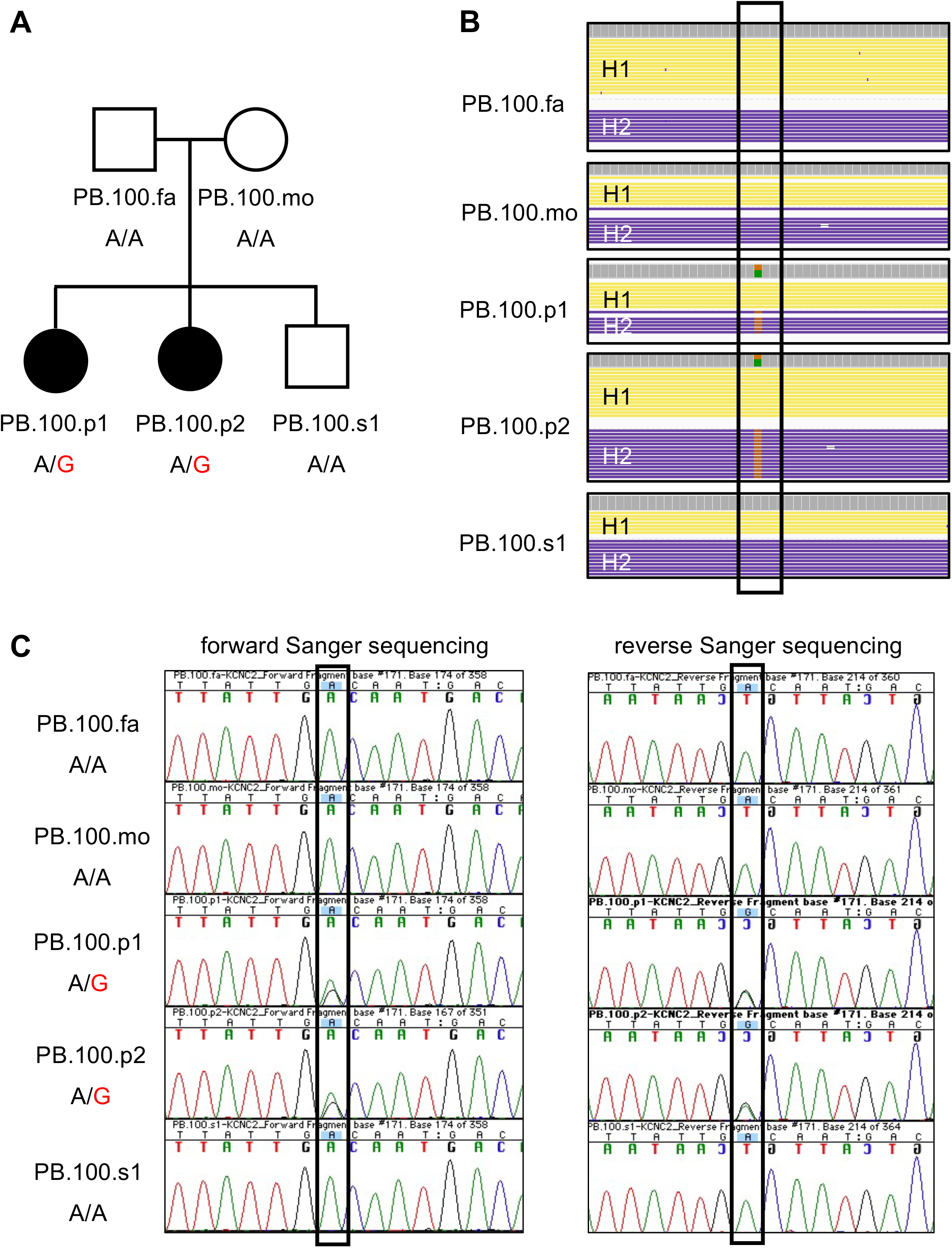
Family assessed in this study and identification of *de novo* missense variant in the *KCNC2* gene. A) Pedigree of family PB.100 with unaffected parents, two female children with autism and epilepsy, and one unaffected male child. Shown below each individual is their genotype for the *KCNC2* variant (chr12:g.75050587A>G). As can be seen, the variant is *de novo* and only seen in the two affected individuals (PB.100.p1, PB.100.p2). B) PacBio read data at and around the *de novo KCNC2* variant position (shown in box). Physically phased read data is shown and is labeled with H1 (haplotype 1) and H2 (haplotype 2) for each individual. The *de novo KCNC2* variant is only identified in the two affected individuals (PB.100.p1, PB.100.p2). C) Sanger confirmation of the *KCNC2* variant detected only in the two affected individuals (PB.100.p1, PB.100.p2). The confirmation is seen in both the forward and reverse Sanger sequencing data.

## RESULTS

### Genomic assessment of PB.100

Three advanced genomic technologies were applied to family PB.100 (Figure 1A) including PacBio HiFi long-read sequencing, 10x Genomics, and Bionano optical mapping to comprehensively characterize genomic variants in all five family members. By focusing on rare (private, inherited) and *de novo* variants only one potentially relevant variant was identified regarding the phenotype of autism in this family; the missense variant in the *KCNC2* gene was identified in both the 10x Genomics and PacBio HiFi technologies. Bionano optical mapping is not able to detect single-nucleotide variants; therefore, it was not possible to identify the variant in the Bionano data.

### Missense variant in KCNC2 gene

Exomiser [Robinson and others 2014] was run in the Genomiser implementation to assess all variants in the genome for potential relevance to the autism phenotype of the affected females. One variant was prioritized as *de novo* in both females with autism but was not present in any other family member (Figure 1B). This variant chr12:g.75050587A>G (human genome build 38) encoded for a missense change of a Valine to Alanine at amino acid 473 (KCNC2:NM_001260497.1:c.1418T>C:p.(Val473Ala)). The Exomiser score was 0.8, the phenotype score was 0.5, and the variant score was 1.0. Several mutation assessment programs rated this variant as severe including a Polyphen2 [Adzhubei and others 2013] damaging score of 0.999, SIFT [Kumar and others 2009] damaging score of 0.010, MutationTaster [Schwarz and others 2014] pathogenic score of 1.000, and a CADD [Kircher and others 2014] score of 26.700. The variant was not present in the gnomAD [Karczewski and others 2020] database and was also not present in our joint-called dataset of 49 publicly available PacBio HiFi genomes. The reference nucleotide and amino acid are completely conserved across all 100 vertebrates underlying the 100-way vertebrate alignment available in the UCSC browser [Kent and others 2002]. Regarding phyloP [Pollard and others 2010] scores, this nucleotide position was in the top 0.05% of all bases in the genome indicating it is an extremely conserved base. We confirmed that the variant was present as *de novo* in both the PacBio and 10x data. Sanger sequencing was also performed on all family members to confirm the variant as *de novo* in the two affected females and was not present in any other family member (Figure 1C).

### Read-backed phasing of the data

Read-backed phasing of the PacBio data was performed using the Whatshap [Martin and others 2016] tool on our DeepVariant [Poplin and others 2018] calls. Through this analysis, a phase-informative single-nucleotide variant was identified 4,880 bp away from the *de novo* variant (Figure 2). The variant was present on the same physical chromosome as the *de novo* variant in both females with autism. The informative variant has been inherited on the paternal chromosome suggesting the *de novo* variant arose as a germline mosaic variant in the paternal germline.

**Figure 2:**
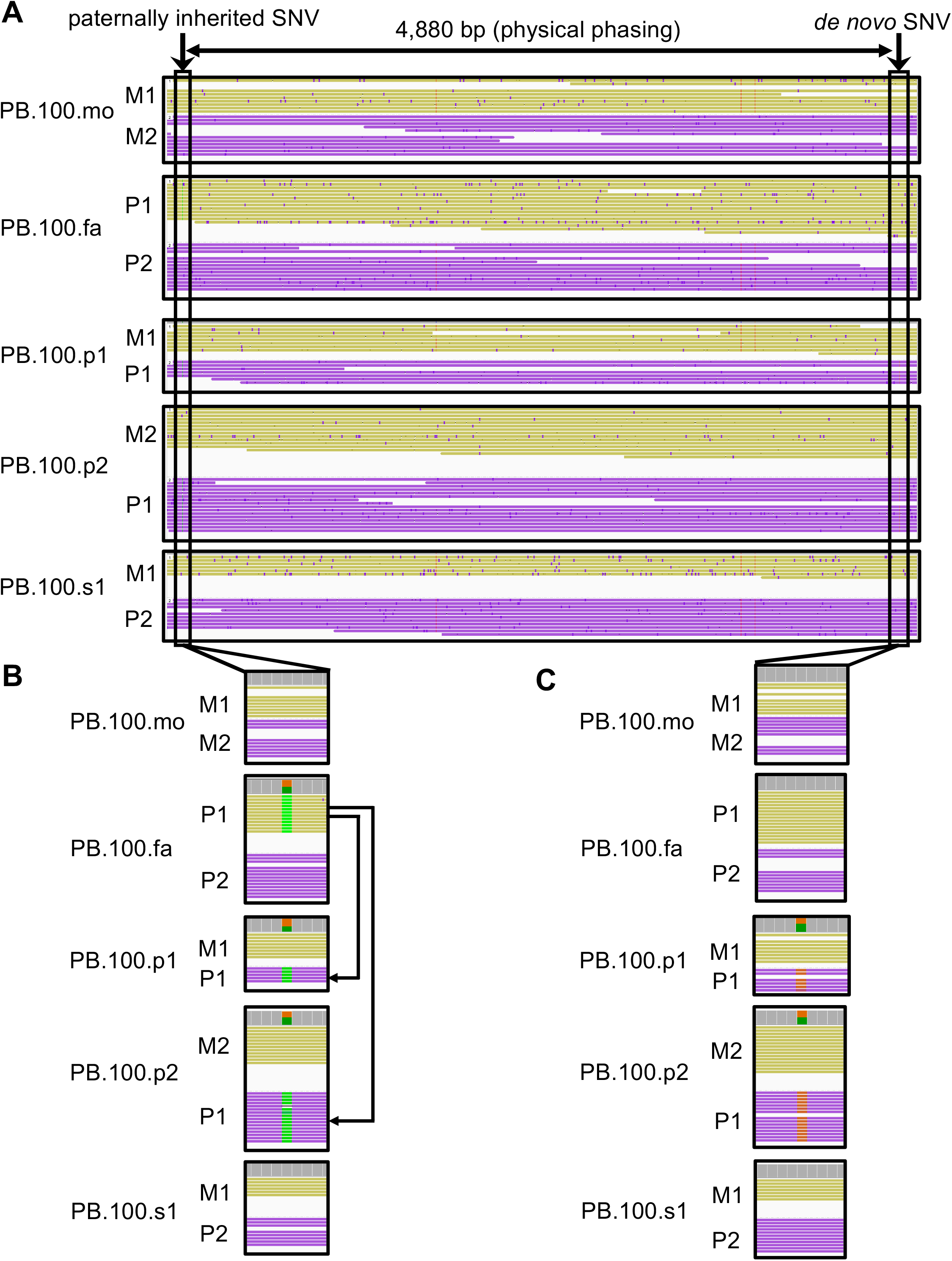
Physical phasing reveals the *de novo* variant arose on the paternal chromosome. A) Shown is an extended window of the phased chromosomes. There is a phase-informative variant (inheritance from paternal chromosome 1 [P1]) 4,880 bp away from the de novo KCNC2 variant and that resides on the same physical chromosome as the *de novo* variant in both PB.100.p1 and PB.100.p2. B) phase by transmission is shown where PB.100.p1 and PB.100.p2 inherit the phase-informative variant from PB.100.fa P1 chromosome. C) the *de novo* variant is shown. Note: chromosome color is based on read-backed phasing result, M1 = maternal chromosome 1, M2 = maternal chromosome 2, P1 = paternal chromosome 1, P2 = paternal chromosome 2.

### Functional consequence of the Val473Ala missense variant

The *KCNC2* gene encodes the Kv3.2 potassium channel. The Kv3.2 potassium channel has 6 transmembrane domains and the Val473Ala missense variant resides at the last residue of the sixth transmembrane domain (S6) near to the intracellular space (Figure 3). This amino acid is highly conserved across orthologous (Figure 4A) and paralogous potassium channels (Figure 4B). In particular, the *exact same amino acid change* was identified in the Kv1.1 protein in a family with episodic ataxia / myokymia syndrome [Browne and others 1994]. Family 1 from that study was identified to have the amino acid change; it was present in all individuals with the syndrome and not in individuals without the syndrome. By functional modeling of both the human Kv1.1 protein and the orthologous Shaker protein in Drosophila it has been shown that *this specific amino acid* change affects the channel function by accelerating current decay, shifting the voltage dependence of activation, and preventing the channel from entering a long-lasting open state [Peters and others 2011].

**Figure 3:**
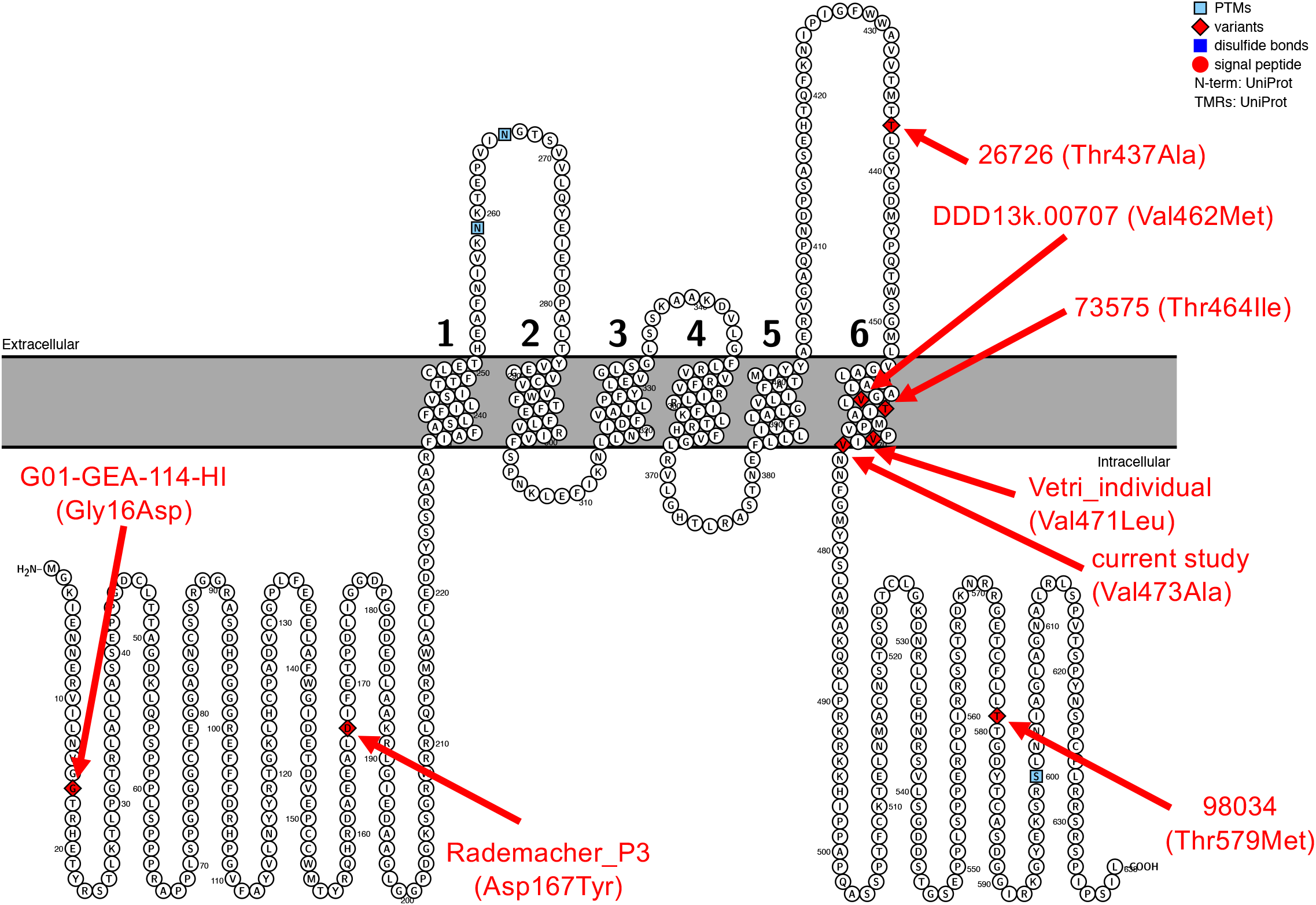
Protein plot of identified Kv3.2 variants in individuals with neurodevelopmental disorders. Shown in red is each variant with sample name from the original publications and the amino acid change. The two individuals (PB.100.p1 and PB.100.p2) are represented by the “current study (Val473Ala)” label. As can be seen, four of the six variants reside in the S6 transmembrane domain of the protein.

**Figure 4:**
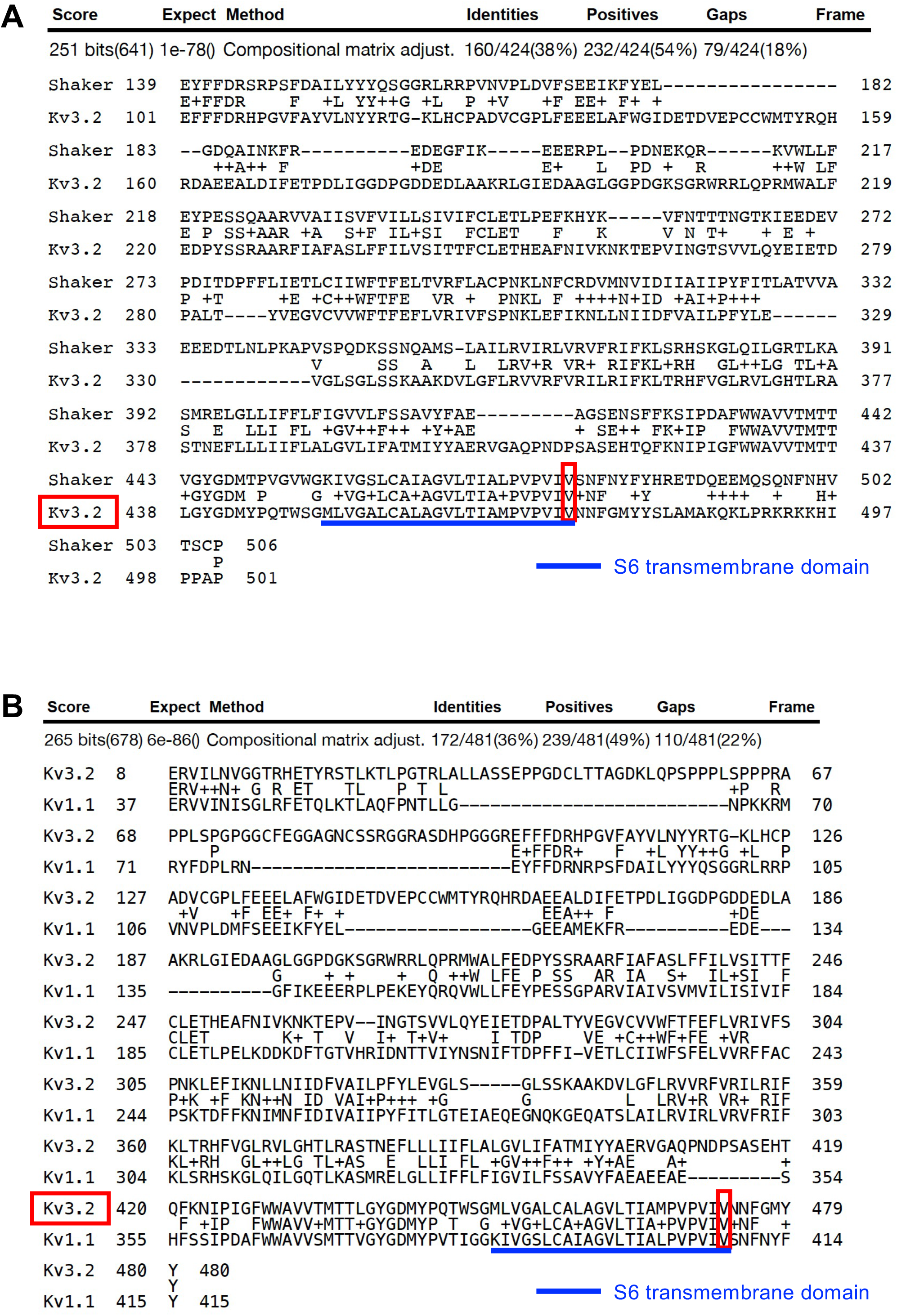
Kv3.2 conservation of the S6 transmembrane domain in the Shaker ortholog and Kv1.1 paralog. A) BLAST of the Drosophila melanogaster Shaker ortholog to the Kv3.2 protein reveals high conservation of the S6 transmembrane domain and complete conservation at the 473 Valine amino acid position. B) BLAST of the human Kv1.1 paralog to the Kv3.2 protein reveals high conservation of the S6 transmembrane domain and complete conservation at the 473 Valine amino acid position. These two proteins were compared since the Valine to Alanine change at the same protein location as in Kv3.2 has already revealed functional consequences of the change in both the Kv1.1 and Shaker proteins.

### Other KCNC2 variants in neurodevelopmental disorders

We searched through the literature and identified 7 additional individuals with neurodevelopmental disorders that had a missense variant in this gene (*de novo* missense p-value = 1.03 × 10^−5^). Four of the variants reside within the S6 transmembrane domain (Figure 3).

### KCNC2 expression

In human tissues, the *KCNC2* gene is expressed highly in the brain and expressed in the pituitary (https://gtexportal.org/home/gene/KCNC2) (Figure 5A). It is lowly expressed in other human tissues. Over the lifetime in the brain, the *KCNC2* gene is expressed most highly after birth and is very highly expressed in the thalamus (https://www.brainspan.org/) (Figure 5B). Further delineation of its expression pattern by single-cell analysis reveals specificity for this channel in inhibitory neurons (https://portal.brain-map.org) (Figure 5C).

**Figure 5:**
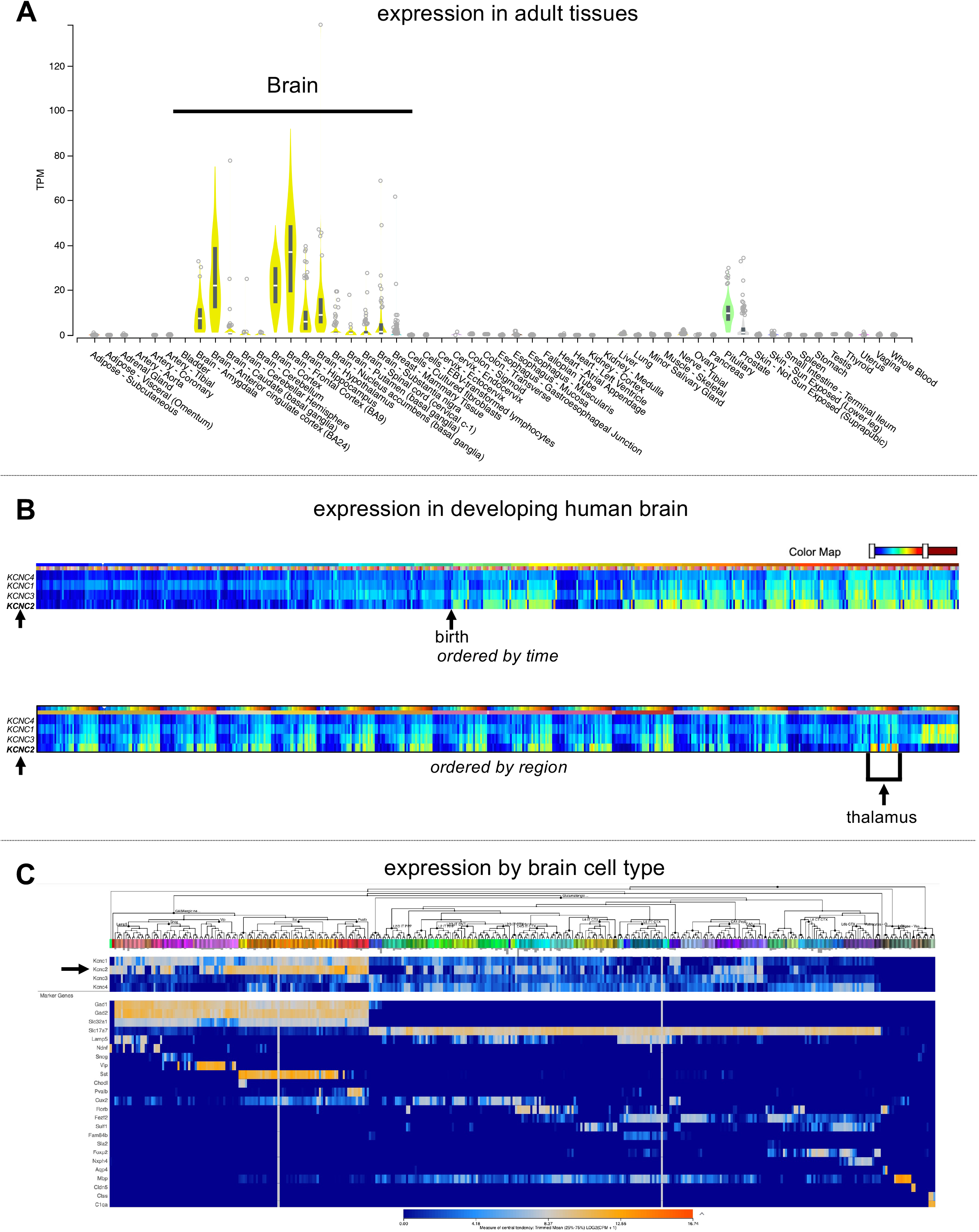
Expression of the *KCNC2* gene. A) The *KCNC2* gene is expressed primarily in the brain from adult human tissues (data from https://gtexportal.org/home/gene/KCNC2). B) In the developing human brain, the *KCNC2* gene is most highly expressed after birth (top) and has the highest expression in the thalamus (bottom) (data from https://www.brainspan.org/). Single-cell expression data reveals the highest expression in GABAergic neurons in the whole cortex and hippocampus from an 8-week-old mouse (data from https://portal.brain-map.org).

## DISCUSSION

In this study, utilization of highly accurate PacBio HiFi long-read sequencing provided genetic answers for a family whose prior clinical genetic test results were negative. By applying a family design of including an unaffected sibling we identified a *de novo* missense variant, phased to the paternal chromosome, in the *KCNC2* gene in both females with autism that was not present in the unaffected sibling. This variant has several effects on the function of the potassium channel. The channel is expressed in GABAergic neurons in the brain with the highest expression in the thalamus. This finding could lead to potential therapeutics regarding the seizure and neurocognitive phenotypes of the affected females.

We hypothesized different possibilities as to why this family was previously unexplained in the clinic and identified several reasons why our approach worked for this family. First, in the original clinical study only one proband was tested by karyotype, array, and whole-exome sequencing. This would mean the *de novo* status of the variant would not have been known and the identification of the variant in the second sibling with autism was also unknown. In contrast, in this study the whole family was sequenced including an unaffected sibling. Second, the *KCNC2* gene is not currently in the list of ACMG genes [Miller and others 2021] even though the variant was predicted pathogenic by all *in silico* analysis tools. We reached out to the diagnostic laboratory of record in July of 2021, and they said that the variant was not called pathogenic because it was not located in an ACMG disease gene. Since identified previous studies had functionally tested *precisely the same missense mutation* in both a human paralog and Drosophila ortholog of this gene, we know that the effect is relevant for the phenotype we see in this family including both autism and epilepsy in both affected individuals. We also note that a recent preprint came out with evidence supporting the role of *KCNC2* in neurodevelopmental disorders [Schwarz and others 2021] and so, it is possible this gene may soon be added to the ACMG list. Finally, long-read sequencing afforded the opportunity to phase the variant and show that in both females with autism the variant arose on the paternal chromosome providing evidence of germline mosaicism.

This study provided genetic answers for this family and serves as a prototypic framework by which long-read sequencing can be utilized in future clinical genomic studies. We recommend that for unexplained families that all family members, including unaffected siblings, be sequenced using long-read sequencing and that both variant detection as well as phasing be performed for each family member. Long-read sequencing enabled “precision genomics” in this case and warrants consideration as a clinical standard, especially in enigmatic cases in which genetic causes are strongly suspected, on our pathway to precision medicine. Finally, we note that careful reassessment of published literature should expand beyond the study of the gene alone. Rather, when there are gene family members, such as the case with potassium channels, those gene family members can also be examined for whether they have similar mutations. This was also a lesson learned in a previous study of mutations in glutamate receptors with regions of high conservation between paralogs [Geisheker and others 2017] and should be more systematically tested in other gene families.

## Data Availability

Data produced in this study are available through dbGaP study phs002698.v1.p1

## ACKNOWLEDGMENTS

Thank you to the family for participating in this study. Thank you to Pacific Biosciences for a SMRT grant to sequence this family. This work was also supported by grants from the National Institutes of Health (R00MH117165, P50HD103525). For Figure 5A, the Genotype-Tissue Expression (GTEx) Project was supported by the Common Fund of the Office of the Director of the National Institutes of Health, and by NCI, NHGRI, NHLBI, NIDA, NIMH, and NINDS. The data used for the analyses described in this manuscript were obtained from: https://gtexportal.org/home/gene/KCNC2 the GTEx Portal on 08/30/21. Data produced in this study are available through dbGaP study phs002698.v1.p1

## REFERENCES

Adzhubei I, Jordan DM, Sunyaev SR. 2013. Predicting functional effect of human missense mutations using PolyPhen-2. Current protocols in human genetics / editorial board, Jonathan L Haines [et al] Chapter 7:Unit7.20.

Altschul SF, Gish W, Miller W, Myers EW, Lipman DJ. 1990. Basic local alignment search tool. J Mol Biol 215(3):403–410.

An JY, Lin K, Zhu L, Werling DM, Dong S, Brand H, Wang HZ, Zhao X, Schwartz GB, Collins RL, Currall BB, Dastmalchi C, Dea J, Duhn C, Gilson MC, Klei L, Liang L, Markenscoff-Papadimitriou E, Pochareddy S, Ahituv N, Buxbaum JD, Coon H, Daly MJ, Kim YS, Marth GT, Neale BM, Quinlan AR, Rubenstein JL, Sestan N, State MW, Willsey AJ, Talkowski ME, Devlin B, Roeder K, Sanders SJ. 2018. Genome-wide de novo risk score implicates promoter variation in autism spectrum disorder. Science (New York, NY) 362(6420).

Browne DL, Gancher ST, Nutt JG, Brunt ER, Smith EA, Kramer P, Litt M. 1994. Episodic ataxia/myokymia syndrome is associated with point mutations in the human potassium channel gene, KCNA1. Nature genetics 8(2):136–140.

Cheng H, Concepcion GT, Feng X, Zhang H, Li H. 2021. Haplotype-resolved de novo assembly using phased assembly graphs with hifiasm. Nature methods.

Coe BP, Stessman HAF, Sulovari A, Geisheker MR, Bakken TE, Lake AM, Dougherty JD, Lein ES, Hormozdiari F, Bernier RA, Eichler EE. 2019. Neurodevelopmental disease genes implicated by de novo mutation and copy number variation morbidity. Nature genetics 51(1):106–116.

Doan RN, Lim ET, De Rubeis S, Betancur C, Cutler DJ, Chiocchetti AG, Overman LM, Soucy A, Goetze S, Freitag CM, Daly MJ, Walsh CA, Buxbaum JD, Yu TW. 2019. Recessive gene disruptions in autism spectrum disorder. Nature genetics 51(7):1092–1098.

Garrison E MG. 2012. Haplotype-based variant detection from short-read sequencing. arXiv preprint 1207.3907 [q-bio.GN]

Gaugler T, Klei L, Sanders SJ, Bodea CA, Goldberg AP, Lee AB, Mahajan M, Manaa D, Pawitan Y, Reichert J, Ripke S, Sandin S. 2014. Most genetic risk for autism resides with common variation. Nature genetics 46(8):881–885.

Geisheker MR, Heymann G, Wang T, Coe BP, Turner TN, Stessman HAF, Hoekzema K, Kvarnung M, Shaw M, Friend K, Liebelt J, Barnett C, Thompson EM, Haan E, Guo H, Anderlid BM, Nordgren A, Lindstrand A, Vandeweyer G, Alberti A, Avola E, Vinci M, Giusto S, Pramparo T, Pierce K, Nalabolu S, Michaelson JJ, Sedlacek Z, Santen GWE, Peeters H, Hakonarson H, Courchesne E, Romano C, Kooy RF, Bernier RA, Nordenskjold M, Gecz J, Xia K, Zweifel LS, Eichler EE. 2017. Hotspots of missense mutation identify neurodevelopmental disorder genes and functional domains. Nature neuroscience 20(8):1043–1051.

Iossifov I, Levy D, Allen J, Ye K, Ronemus M, Lee Y-h, Yamrom B, Wigler M. 2015. Low load for disruptive mutations in autism genes and their biased transmission. Proceedings of the National Academy of Sciences 112(41):E5600–E5607.

Iossifov I, O’Roak BJ, Sanders SJ, Ronemus M, Krumm N, Levy D, Stessman HA, Witherspoon KT, Vives L, Patterson KE, Smith JD, Paeper B, Nickerson DA, Dea J, Dong S, Gonzalez LE, Mandell JD, Mane SM, Murtha MT, Sullivan CA, Walker MF, Waqar Z, Wei L, Willsey AJ, Yamrom B, Lee Y-h, Grabowska E, Dalkic E, Wang Z, Marks S, Andrews P, Leotta A, Kendall J, Hakker I, Rosenbaum J, Ma B, Rodgers L, Troge J, Narzisi G, Yoon S, Schatz MC, Ye K, McCombie WR, Shendure J, Eichler EE, State MW, Wigler M. 2014. The contribution of de novo coding mutations to autism spectrum disorder. Nature.

Jacquemont S, Coe BP, Hersch M, Duyzend MH, Krumm N, Bergmann S, Beckmann JS, Rosenfeld JA, Eichler EE. 2014. A higher mutational burden in females supports a “female protective model” in neurodevelopmental disorders. American journal of human genetics 94(3):415–425.

Kaplanis J, Samocha KE, Wiel L, Zhang Z, Arvai KJ, Eberhardt RY, Gallone G, Lelieveld SH, Martin HC, McRae JF, Short PJ, Torene RI, de Boer E, Danecek P, Gardner EJ, Huang N, Lord J, Martincorena I, Pfundt R, Reijnders MRF, Yeung A, Yntema HG, Vissers L, Juusola J, Wright CF, Brunner HG, Firth HV, FitzPatrick DR, Barrett JC, Hurles ME, Gilissen C, Retterer K. 2020. Evidence for 28 genetic disorders discovered by combining healthcare and research data. Nature 586(7831):757–762.

Karczewski KJ, Francioli LC, Tiao G, Cummings BB, Alföldi J, Wang Q, Collins RL, Laricchia KM, Ganna A, Birnbaum DP, Gauthier LD, Brand H, Solomonson M, Watts NA, Rhodes D, Singer-Berk M, England EM, Seaby EG, Kosmicki JA, Walters RK, Tashman K, Farjoun Y, Banks E, Poterba T, Wang A, Seed C, Whiffin N, Chong JX, Samocha KE, Pierce-Hoffman E, Zappala Z, O’Donnell-Luria AH, Minikel EV, Weisburd B, Lek M, Ware JS, Vittal C, Armean IM, Bergelson L, Cibulskis K, Connolly KM, Covarrubias M, Donnelly S, Ferriera S, Gabriel S, Gentry J, Gupta N, Jeandet T, Kaplan D, Llanwarne C, Munshi R, Novod S, Petrillo N, Roazen D, Ruano-Rubio V, Saltzman A, Schleicher M, Soto J, Tibbetts K, Tolonen C, Wade G, Talkowski ME, Neale BM, Daly MJ, MacArthur DG. 2020. The mutational constraint spectrum quantified from variation in 141,456 humans. Nature 581(7809):434–443.

Kent WJ, Sugnet CW, Furey TS, Roskin KM, Pringle TH, Zahler AM, Haussler D. 2002. The human genome browser at UCSC. Genome research 12(6):996–1006.

Kircher M, Witten DM, Jain P, O’Roak BJ, Cooper GM, Shendure J. 2014. A general framework for estimating the relative pathogenicity of human genetic variants. Nature genetics 46(3):310–315.

Krumm N, Turner TN, Baker C, Vives L, Mohajeri K, Witherspoon K, Raja A, Coe BP, Stessman HA, He ZX, Leal SM, Bernier R, Eichler EE. 2015. Excess of rare, inherited truncating mutations in autism. Nature genetics 47(6):582–588.

Kumar P, Henikoff S, Ng PC. 2009. Predicting the effects of coding non-synonymous variants on protein function using the SIFT algorithm. Nature protocols 4(7):1073–1081.

Levy D, Ronemus M, Yamrom B, Lee YH, Leotta A, Kendall J, Marks S, Lakshmi B, Pai D, Ye K, Buja A, Krieger A, Yoon S, Troge J, Rodgers L, Iossifov I, Wigler M. 2011. Rare de novo and transmitted copy-number variation in autistic spectrum disorders. Neuron 70(5):886–897.

Martin M, Patterson M, Garg S O, Fischer S, Pisanti N, Klau GW, Schöenhuth A, Marschall T. 2016. WhatsHap: fast and accurate read-based phasing. bioRxiv:085050.

Miller DT, Lee K, Chung WK, Gordon AS, Herman GE, Klein TE, Stewart DR, Amendola LM, Adelman K, Bale SJ, Gollob MH, Harrison SM, Hershberger RE, McKelvey K, Richards CS, Vlangos CN, Watson MS, Martin CL. 2021. ACMG SF v3.0 list for reporting of secondary findings in clinical exome and genome sequencing: a policy statement of the American College of Medical Genetics and Genomics (ACMG). Genetics in medicine : official journal of the American College of Medical Genetics 23(8):1381–1390.

Neale BM, Kou Y, Liu L, Ma’ayan A, Samocha KE, Sabo A, Lin CF, Stevens C, Wang LS, Makarov V, Polak P, Yoon S, Maguire J, Crawford EL, Campbell NG, Geller ET, Valladares O, Schafer C, Liu H, Zhao T, Cai G, Lihm J, Dannenfelser R, Jabado O, Peralta Z, Nagaswamy U, Muzny D, Reid JG, Newsham I, Wu Y, Lewis L, Han Y, Voight BF, Lim E, Rossin E, Kirby A, Flannick J, Fromer M, Shakir K, Fennell T, Garimella K, Banks E, Poplin R, Gabriel S, DePristo M, Wimbish JR, Boone BE, Levy SE, Betancur C, Sunyaev S, Boerwinkle E, Buxbaum JD, Cook EH, Jr., Devlin B, Gibbs RA, Roeder K, Schellenberg GD, Sutcliffe JS, Daly MJ. 2012. Patterns and rates of exonic de novo mutations in autism spectrum disorders. Nature 485(7397):242–245.

Nurk S, Walenz BP, Rhie A, Vollger MR, Logsdon GA, Grothe R, Miga KH, Eichler EE, Phillippy AM, Koren S. 2020. HiCanu: accurate assembly of segmental duplications, satellites, and allelic variants from high-fidelity long reads. Genome research 30(9):1291–1305.

Omasits U, Ahrens CH, Müller S, Wollscheid B. 2013. Protter: interactive protein feature visualization and integration with experimental proteomic data. Bioinformatics (Oxford, England) 30(6):884–886.

Padhi EM, Hayeck TJ, Cheng Z, Chatterjee S, Mannion BJ, Byrska-Bishop M, Willems M, Pinson L, Redon S, Benech C, Uguen K, Audebert-Bellanger S, Le Marechal C, Férec C, Efthymiou S, Rahman F, Maqbool S, Maroofian R, Houlden H, Musunuri R, Narzisi G, Abhyankar A, Hunter RD, Akiyama J, Fries LE, Ng JK, Mehinovic E, Stong N, Allen AS, Dickel DE, Bernier RA, Gorkin DU, Pennacchio LA, Zody MC, Turner TN. 2021. Coding and noncoding variants in EBF3 are involved in HADDS and simplex autism. Hum Genomics 15(1):44.

Peters CJ, Werry D, Gill HS, Accili EA, Fedida D. 2011. Mechanism of accelerated current decay caused by an episodic ataxia type-1-associated mutant in a potassium channel pore. The Journal of neuroscience : the official journal of the Society for Neuroscience 31(48):17449–17459.

Pollard KS, Hubisz MJ, Rosenbloom KR, Siepel A. 2010. Detection of nonneutral substitution rates on mammalian phylogenies. Genome research 20(1):110–121.

Poplin R, Chang PC, Alexander D, Schwartz S, Colthurst T, Ku A, Newburger D, Dijamco J, Nguyen N, Afshar PT, Gross SS, Dorfman L, McLean CY, DePristo MA. 2018. A universal SNP and small-indel variant caller using deep neural networks. Nature biotechnology 36(10):983–987.

Purcell S, Neale B, Todd-Brown K, Thomas L, Ferreira MA, Bender D, Maller J, Sklar P, de Bakker PI, Daly MJ, Sham PC. 2007. PLINK: a tool set for whole-genome association and population-based linkage analyses. American journal of human genetics 81(3):559–575.

Rademacher A, Schwarz N, Seiffert S, Pendziwiat M, Rohr A, van Baalen A, Helbig I, Weber Y, Muhle H. 2020. Whole-Exome Sequencing in NF1-Related West Syndrome Leads to the Identification of KCNC2 as a Novel Candidate Gene for Epilepsy. Neuropediatrics 51(5):368–372.

Robinson PN, Köhler S, Oellrich A, Wang K, Mungall CJ, Lewis SE, Washington N, Bauer S, Seelow D, Krawitz P, Gilissen C, Haendel M, Smedley D. 2014. Improved exome prioritization of disease genes through cross-species phenotype comparison. Genome research 24(2):340–348.

Samocha KE, Robinson EB, Sanders SJ, Stevens C, Sabo A, McGrath LM, Kosmicki JA, Rehnstrom K, Mallick S, Kirby A, Wall DP, MacArthur DG, Gabriel SB, DePristo M, Purcell SM, Palotie A, Boerwinkle E, Buxbaum JD, Cook EH, Jr., Gibbs RA, Schellenberg GD, Sutcliffe JS, Devlin B, Roeder K, Neale BM, Daly MJ. 2014. A framework for the interpretation of de novo mutation in human disease. Nature genetics 46(9):944–950.

Sanders SJ, He X, Willsey AJ, Ercan-Sencicek AG, Samocha KE, Cicek AE, Murtha MT, Bal VH, Bishop SL, Dong S, Goldberg AP, Jinlu C, Keaney JF, 3rd, Klei L, Mandell JD, Moreno-De-Luca D, Poultney CS, Robinson EB, Smith L, Solli-Nowlan T, Su MY, Teran NA, Walker MF, Werling DM, Beaudet AL, Cantor RM, Fombonne E, Geschwind DH, Grice DE, Lord C, Lowe JK, Mane SM, Martin DM, Morrow EM, Talkowski ME, Sutcliffe JS, Walsh CA, Yu TW, Ledbetter DH, Martin CL, Cook EH, Buxbaum JD, Daly MJ, Devlin B, Roeder K, State MW. 2015. Insights into Autism Spectrum Disorder Genomic Architecture and Biology from 71 Risk Loci. Neuron 87(6):1215–1233.

Satterstrom FK, Kosmicki JA, Wang J, Breen MS, De Rubeis S, An JY, Peng M, Collins R, Grove J, Klei L, Stevens C, Reichert J, Mulhern MS, Artomov M, Gerges S, Sheppard B, Xu X, Bhaduri A, Norman U, Brand H, Schwartz G, Nguyen R, Guerrero EE, Dias C, Betancur C, Cook EH, Gallagher L, Gill M, Sutcliffe JS, Thurm A, Zwick ME, Børglum AD, State MW, Cicek AE, Talkowski ME, Cutler DJ, Devlin B, Sanders SJ, Roeder K, Daly MJ, Buxbaum JD. 2020. Large-Scale Exome Sequencing Study Implicates Both Developmental and Functional Changes in the Neurobiology of Autism. Cell 180(3):568-584.e523.

Schwarz JM, Cooper DN, Schuelke M, Seelow D. 2014. MutationTaster2: mutation prediction for the deep-sequencing age. Nature methods 11(4):361–362.

Schwarz N, Seiffert S, Pendziwiat M, Rademacher A, Brünger T, Hedrich UBS, Augustijn PB, Baier H, Bayat A, Bisulli F, Buono RJ, Bruria BZ, Doyle MG, Guerrini R, Heimer G, Iacomino M, Kearney H, Klein KM, Kousiappa I, Kunz WS, Lerche H, Licchetta L, Lohmann E, Minardi R, McDonald M, Montgomery S, Mulahasanovic L, Oegema R, Ortal B, Papacostas SS, Ragona F, Granata T, Reif PS, Rosenow F, Rothschild A, Scudieri P, Striano P, Tinuper P, Tanteles GA, Vetro A, Zahnert F, Zara F, Lal D, May P, Muhle H, Helbig I, Weber Y. 2021. Heterozygous variants in <em>KCNC2</em> cause a broad spectrum of epilepsy phenotypes associated with characteristic functional alterations. medRxiv:2021.2005.2021.21257099.

Turner TN, Coe BP, Dickel DE, Hoekzema K, Nelson BJ, Zody MC, Kronenberg ZN, Hormozdiari F, Raja A, Pennacchio LA, Darnell RB, Eichler EE. 2017. Genomic Patterns of De Novo Mutation in Simplex Autism. Cell 171(3):710-722.e712.

Turner TN, Hormozdiari F, Duyzend MH, McClymont SA, Hook PW, Iossifov I, Raja A, Baker C, Hoekzema K, Stessman HA, Zody MC, Nelson BJ, Huddleston J, Sandstrom R, Smith JD, Hanna D, Swanson JM, Faustman EM, Bamshad MJ, Stamatoyannopoulos J, Nickerson DA, McCallion AS, Darnell R, Eichler EE. 2016. Genome Sequencing of Autism-Affected Families Reveals Disruption of Putative Noncoding Regulatory DNA. American journal of human genetics 98(1):58–74.

Turner TN, Sharma K, Oh EC, Liu YP, Collins RL, Sosa MX, Auer DR, Brand H, Sanders SJ, Moreno-De-Luca D, Pihur V, Plona T, Pike K, Soppet DR, Smith MW, Cheung SW, Martin CL, State MW, Talkowski ME, Cook E, Huganir R, Katsanis N, Chakravarti A. 2015. Loss of δ-catenin function in severe autism. Nature 520(7545):51–56.

Turner TN, Wilfert AB, Bakken TE, Bernier RA, Pepper MR, Zhang Z, Torene RI, Retterer K, Eichler EE. 2019. Sex-Based Analysis of De Novo Variants in Neurodevelopmental Disorders. American journal of human genetics 105(6):1274–1285.

Vetri L, Calì F, Vinci M, Amato C, Roccella M, Granata T, Freri E, Solazzi R, Romano V, Elia M. 2020. A de novo heterozygous mutation in KCNC2 gene implicated in severe developmental and epileptic encephalopathy. Eur J Med Genet 63(4):103848.

Ware JS, Samocha KE, Homsy J, Daly MJ. 2015. Interpreting de novo variation in human disease using denovolyzeR. Current protocols in human genetics 87:7.25.21-15.

Weiner DJ, Wigdor EM, Ripke S, Walters RK, Kosmicki JA, Grove J, Samocha KE, Goldstein JI, Okbay A, Bybjerg-Grauholm J, Werge T, Hougaard DM, Taylor J, Skuse D, Devlin B, Anney R, Sanders SJ, Bishop S, Mortensen PB, Børglum AD, Smith GD, Daly MJ, Robinson EB. 2017. Polygenic transmission disequilibrium confirms that common and rare variation act additively to create risk for autism spectrum disorders. Nature genetics 49(7):978–985.

Wenger AM, Peluso P, Rowell WJ, Chang PC, Hall RJ, Concepcion GT, Ebler J, Fungtammasan A, Kolesnikov A, Olson ND, Töpfer A, Alonge M, Mahmoud M, Qian Y, Chin CS, Phillippy AM, Schatz MC, Myers G, DePristo MA, Ruan J, Marschall T, Sedlazeck FJ, Zook JM, Li H, Koren S, Carroll A, Rank DR, Hunkapiller MW. 2019. Accurate circular consensus long-read sequencing improves variant detection and assembly of a human genome. Nature biotechnology 37(10):1155–1162.

Wilfert AB, Turner TN, Murali SC, Hsieh P, Sulovari A, Wang T, Coe BP, Guo H, Hoekzema K, Bakken TE, Winterkorn LH, Evani US, Byrska-Bishop M, Earl RK, Bernier RA, Zody MC, Eichler EE. 2021. Recent ultra-rare inherited variants implicate new autism candidate risk genes. Nature genetics.

Yun T, Li H, Chang P-C, Lin MF, Carroll A, McLean CY. 2020. Accurate, scalable cohort variant calls using DeepVariant and GLnexus. bioRxiv:2020.2002.2010.942086.

Zhou J, Park CY, Theesfeld CL, Wong AK, Yuan Y, Scheckel C, Fak JJ, Funk J, Yao K, Tajima Y, Packer A, Darnell RB, Troyanskaya OG. 2019. Whole-genome deep-learning analysis identifies contribution of noncoding mutations to autism risk. Nature genetics 51(6):973–980.

